# In-house assembled protective devices in laboratory safety against SARS-nCoV-2 in clinical biochemistry laboratory of a COVID dedicated hospital

**DOI:** 10.1101/2020.08.24.20155713

**Authors:** Abhishek Dubey, Aastha Bansal, Subash Chandra Sonkar, Binita Goswami, Naina Makwane, Vikas Manchanda, Bidhan Chandra Koner

## Abstract

Health Care Workers (HCWs) of diagnostic laboratory handling COVID positive samples are at risk and need to take protective measures. Many protective materials were not available when the pandemic reached India forcing laboratory managers to take innovative measures to protect the laboratory staffs. We made face shields from OHP sheets and substitute of biosafety cabinets from cardboard boxes fitted with hypochlorite spraying devices. Here we present if these two in-house developed safety devices when incorporated in standard operating procedure (SOP) of laboratory safety were effective in clinical biochemistry laboratory of dedicated COVID hospitals. We assessed contamination of laboratory surfaces (n=6) and rate of SARS-nCov-2 positivity from their nasal and throat swab by RT-PCR among laboratory personnel (n=18) after 14 days of their use along with other routine safety devices like use of gloves, surgical masks, OT gowns etc. These HCWs were checked regularly for signs and symptoms of COVID-19 and none had any signs and symptoms during these 14days. The SARS-nCov-2 test report was negative for the staff members and no surface contamination was detected. We conclude that innovative and cost effective protective devices can be built in-house with locally available resources and are effective in preventing the spread of COVID 19 among the staff working in clinical biochemistry laboratories. Laboratory managers in resource scarce areas need to be innovative to face such sudden safety challenges like COVID-19 pandemic.

**The highlight of the manuscript are:**

- Strengthening the Basics Approaches to protect the lab personnel in dedicated COVID hospital of Low-Resource Settings.
- Designed and developed in-house standard operating procedure (SOP) to fill the gap and evaluate the effect in dedicated COVID-19 hospitals.
- Innovative protective devices made from OHP sheets and cardboard boxes fitted with hypochlorite spraying devices as alternatives to biosafety cabinets on contamination of laboratory surfaces.
- Performance of the devices were clinically validated and it can be used as alternative in low resources settings.

## Introduction

Last day of the year of 2019 (December 31^st^ 2019) was the day when the first Coronavirus case was identified and came into light officially in China. As on date, 2nd June 2020, after five months, there is a lot of fear and anxiety about this pandemic with drastic developments in the worldwide healthcare scenario. According to World Health Organization (WHO) reports, there have been 6,194,533 confirmed cases of COVID-19, including 376,320 deaths. As per the reported cases USA and Europe are the top two continents where COVID-19 confirmed cases are highest in the world with 2,905,432 and 2,175,941 respectively. In India, as per Indian Council of Medical Research (ICMR) New Delhi, as on date; India with 97581 active Cases, 95526 Cured / Discharged cases and 5598 Deaths reported till date.

In a low resource settings nation like India with a population of 1.34 billion, the existing healthcare infrastructure and delivery system have already been facing challenges of accessibility and affordability. Unfortunately, this COVID-19 pandemic further increased the already overburdened HCWs. The doctors, nurses, paramedics, and other hospital staff are bearing the onslaught of this infection directly. The number of patients of COVID-19 and their contacts undergoing quarantine is increasing by the day, and these will continue to increase at a faster rate in the days to come because of asymptomatic and community spreading. In this difficult scenario, HCWs are undoubtedly fighting with all their strength. These healthcare personnel are under tremendous physical and mental stress. Apart from their health, they are concerned about transmitting the infection back to their homes, to their families and friends.

The HCWs who are at the forefront of managing the pandemic are at risk of contracting the disease as it is highly contagious in nature. Besides the treating physicians and the intensive visits who are coming in direct contact with the patients, laboratory personnel working in clinical biochemistry laboratories are involved in carrying out the various diagnostic tests required for management of these COVID 19 patients^1^. Many such patients need to be tested for routine parameters like plasma glucose, liver function tests, kidney function tests, etc. The critical patients often need specialized tests like C Reactive protein, ferritin, Procalcitonin, D dimer etc. The risk of exposure of laboratory personnel to virus is substantial in view of presence of viral load although it is lesser than that of treating clinicians and interventionists^2^.

SARS-CoV-2 causing COVID-19 is an RNA virus and studies have confirmed its presence in respiratory tract samples, gastro-intestinal tract samples, feces (25-30%) and blood (1-15%) ^3-4^. The virus also spreads through contaminated fomites and surfaces which necessitate sanitizing all surfaces and fomites like centrifuge machines, bench tops, switches, elevator buttons, door handles etc ^5^. However, the major route of transmission of COVID-19 is through respiratory route and that mostly occurs through droplets produced during coughing and sneezing^6^. Many laboratory procedures like centrifugation of samples, decapping of sample tubes, pipetting etc. also generate aerosols that are potential sources of infection ^7^. Various measures are recommended by CDC and WHO for preventing such spread of COVID-19 among laboratory personnel ^8^. Use of gloves, face masks, face shield, PPE, biosafety cabinet for aerosol generating procedure, decontamination of surfaces by hypochlorite or other solutions, proper disposal of biomedical wastes generated in diagnostic laboratories are a few of them ^9^. There is sudden huge demand and an immense resource crunch on the already shoestring healthcare budget of developing countries leading to shortage of PPE, face shields, masks, caps, sanitizers etc. due to the surge in the number of COVID-19 cases. The laboratory from where we are sharing the experience did not get PPEs, face shields and biosafety cabinets on initial days when samples from COVID 19 patients started coming. It took some time to arrange these equipment. Disposable face shields were made in-house from OHP sheets. Cardboard boxes fitted with saran wrap on its upper surface, a device to spray hypochlorite and two holes to introduce gloved hands inside, those get closed on taking out the hand was made in-house and was used as substitute of biosafety cabinet for decapping of sample tubes and pipetting of samples. Strict use of OT gowns by laboratory staff when on duty and their daily cleaning with soapy water was ensured besides other standard precautions like use of gloves, surgical masks, regular surface decontamination of working bench and lab instruments and biomedical waste disposal^10^. Keeping the safety of laboratory personnel paramount and taking cognizance of the limited resources available, we devised these low cost alternatives to ensure safety while handling samples in our lab. The present study presents the effectiveness of SOP using these innovative and cost effective protective devices built in-house with limited resources in preventing contamination of laboratory working surfaces and equipment and in preventing the spread of COVID-19 among the HCWs working in clinical biochemistry laboratories.

## Materials and Methods

The experience is shared from the clinical Biochemistry laboratory of LN Hospital, New Delhi, India that has been ear-marked as an exclusive facility catering only to COVID-19 patients.

The SOP developed was a part of the managerial and operational effort when lab personnel suddenly reported of arrival of blood samples in the lab for investigations from patients suffering from COVID-19 and the results presented in this article are by product of the activities done during this process on subsequent days till the lab could ensure standard safety measures recommended for such labs ^11-16^.

A three member team was formed immediately to make a SOP for Biochemistry lab processing COVID samples based on WHO/CDC guideline, to find out the gaps/deficits in lab for implementing those standard biosafety recommendations and to recommend the best possible measures to tackle the gaps^8^.

The SOP was followed for 14 days till a class II bio-safety cabinet and recommended PPEs were made available. During this period the staff members were instructed to report any symptoms such as sore throat, cough, fever, diarrhea, loss of taste or smell or any other symptoms during their posting in the lab. COVID-19 screening facility by RT-PCR as recommended by ICMR could not be arranged for lab staff on 6^th^ day as COVID-19 diagnostic facility had limited capacity and was overloaded with a number of samples from suspected cases. The staff members underwent COVID-19 testing on 14^th^ day. Both nasal and throat swabs were sent to the virology laboratory in the Microbiology department for RT-PCR detection of the virus causing COVID-19. One technician refused to provide a sample because he was fasting during Ramdan and as per his faith the introduction of a swab to the nose or throat is equivalent to breaking his fasting. His faith was respected. Swabs from lab surfaces from sample collection and processing area, centrifuge machine, cardboard box used for decapping and sample loading area of clinical chemistry analyzer were sent for viral detection by RT-PCR to assess surface contamination.

### Detection of virus causing COVID-19 by RT-PCR

Total RNA was isolated from fresh Throat & Nasal swab (TS/NS-200 ul/ Specimens) collected from laboratory staff and followed by MagGenome Xpress RNA Isolation Kit as per instruction manuals ^18^. The surface swabs were also processed in a similar way. The quality and quantity of RNA were estimated by using NanoDrop ND-1000 spectrophotometer (Thermo Fisher Scientific, Waltham, MA, USA) and extracted RNA was stored at -20° C for further use. Quality of isolated total RNA from all samples was checked by RNaseP gene amplification which was used as internal control. Combined reverse transcription of viral RNA and PCR amplification using real-time reverse transcriptase PCR (RT-PCR) methods was done as per instruction manuals ^19-20^.

RT-PCR amplification was carried out in 25μl reaction mix containing 12.5μl of 2X Reaction mix, 1.5μl of target specific primers and Probe (Egene, RNasep gene, RDRP gene and Orf Gene), and 5μl of total RNA isolated from clinical sample, 1.0μl AgPath One-Step RT-PCR enzymes (Applied Biosystem). Each set of RT-PCR assays included a negative control (nucleases free water instead of RNA sample) and a positive control of COVID-19. Amplification of COVID-19 was performed in the thermal cycler (Applied Biosystems QuantStudio 5 Real-Time PCR System) using the following conditions; Reverse Transcriptions 55°C for 30min, Taq inhibitor inactivation 95 °C for 3 min, PCR Amplifications; 45 cycles of 95 °C for 15s, 58 °C for 30 s Data Collections) as per published protocol ^21^. In this one step RT PCR assays, a set of primer and probe used are shown in table No.2.

## Results

The residents (n=5) and lab technicians (n=10) did 4 to 5 days lab duty of 12 hour duration during this 14days period. Each nursing orderlies did duty for 8 to 10 days in 8-12 hr shifts during this period. None of the staff members reported sick during this period. The nasal and throat swab test for virus causing COVID-19 of all technical staff (n=10), residents (n=5) and nursing orderlies (n=3) also came out to be negative. No surface contamination with the virus of the work area and instruments was detected.

## Discussion

This article presents the data that evaluated the effectiveness of a SOP that used a face shield made in-house from OHP sheets and innovative device made from card board fitted with hypochlorite spraying device as alternative to biosafety cabinet along with other standard biosafety measures in preventing spread of COVID-19 virus among laboratory personnel working in biochemistry laboratory in a dedicated COVID-19 hospital. The data shows that at the end of 14days of following the SOP, none of the lab personnel including those who did high risk jobs that produce aerosols developed signs and symptoms of COVID-19 infection and RT-PCR based COVID-19 testing was negative for all lab staff. This indicates that these low cost, innovative devices built in-house were effective in preventing COVID-19 spread through aerosol among lab staff. We attribute this prevention to use of these innovative devices along with other protective gears like face mask, gloves etc. However, we do not claim that it is solely due to our innovative devices but is contributed by these devices also. However, we could not keep a control arm in our study to prove our claim. Another limitation is that most of the COVID-19 cases are asymptomatic and hence, clinical monitoring by self-reporting of symptoms is bound to miss the diagnosis in most of the cases. Even diagnosis by RT-PCR is only 70-80% sensitive ^22^.

Even the lab surfaces and equipment did not show the presence of the virus by RT-PCR testing of the swab taken from the working surfaces and equipment. So it proves that transmission through surface contamination did not occur. The Substitution of a biosafety cabinet with an innovative cardboard box might have acted as a barrier for spread of aerosol although it was not having a negative pressure. However, regular decontamination of the working surfaces with hypochlorite and instruments with 70% ethanol might be a major contributor of prevention of surface contamination. Chance of survival of the virus inside the cardboard box was prevented by spray of hypochlorite solution after each session of decapping and pipetting. This spraying device also helped us use the device for 1-2days.

We conclude that in resource scarce health set up where standard CDC or WHO recommended biosafety measures cannot be totally followed, such simple, low cost and innovative devices made in-house or locally from locally available materials can be effectively used in such sudden outbreak of contagious viral infections. With understanding of the basic principle of barrier method, innovations are possible and are key to success in such critical periods.

## Data Availability

N/A

## Contributorship

BCK and BG researched literature and conceived the study, Supervision, SOPs design and Review & Editing Manuscript. AD and AB: collected sample, executed experiments as per SOPs, Writing – Original Draft and Data Analysis. SCS Conducted RT-PCR based wet labs experiments, Data Analysis, Writing – Original Draft, Review & Editing, VM: Review & Editing, Supervision and for providing his lab for RT-PCR experiments. MN: Review & Editing and Supervision during SOPS design and executions. All authors reviewed and edited the manuscript and approved the final version of the manuscript

## Acknowledgement

The authors also acknowledge the Department of Biochemistry, Department of Microbiology and Multidisciplinary Research Unit, Maulana Azad Medical College & Associated Hospitals, New Delhi- 110002, INDIA for necessary facilities in design and implementation of experiments including preparation of the manuscript.

---

**Fig-1:**
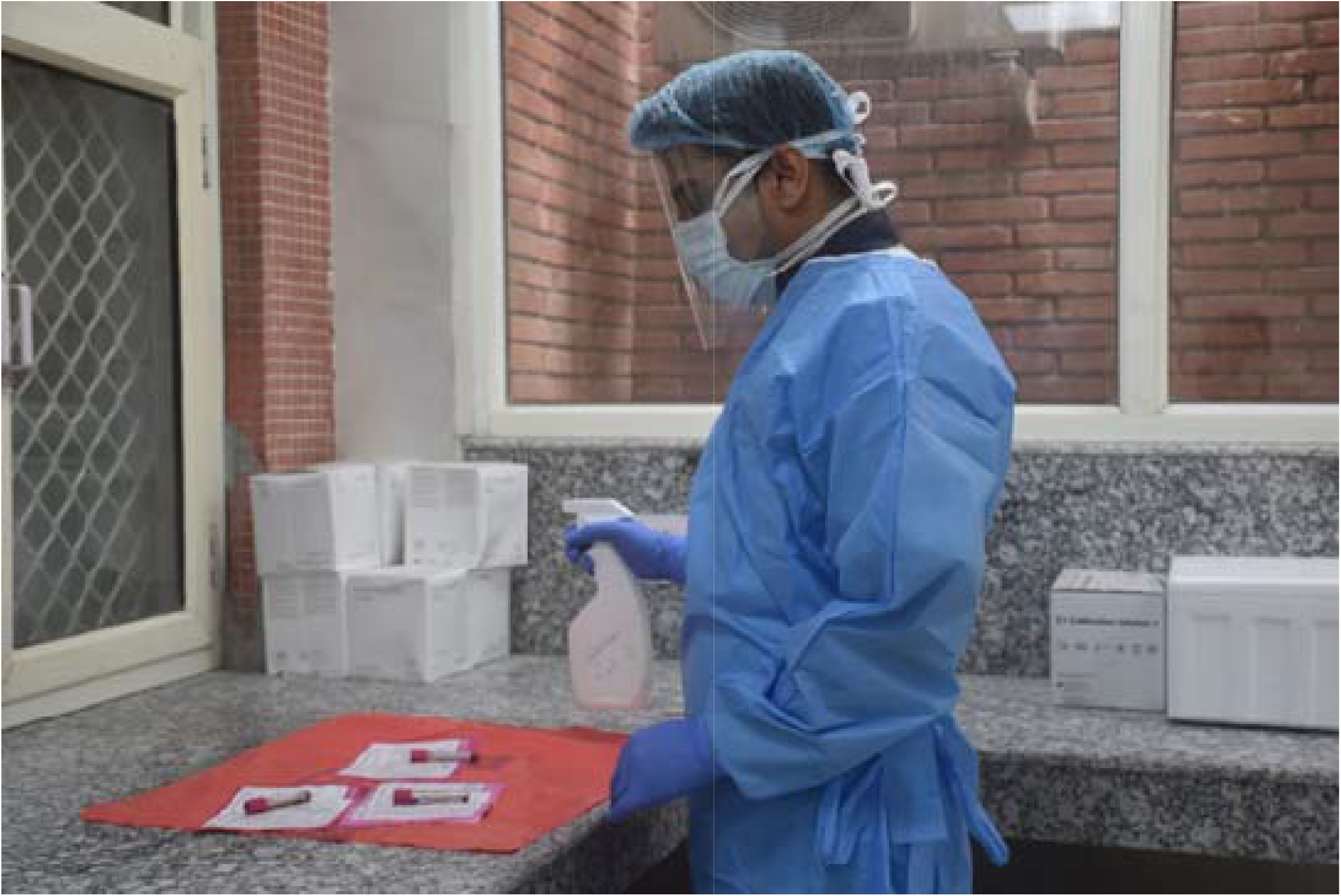
**Spraying of hypochlorite on the samples and the requisition slips**

**Fig 2:**
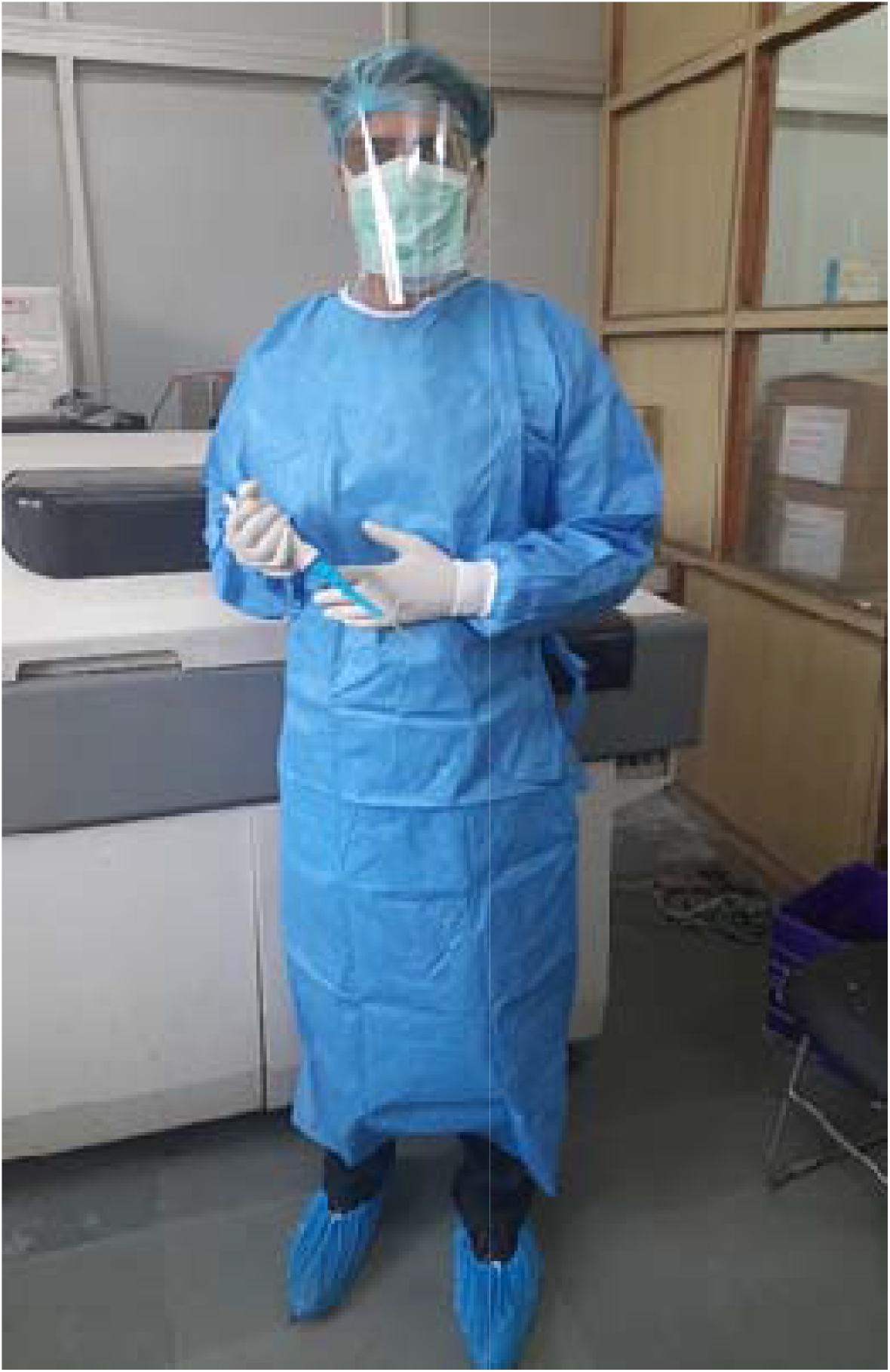
A technician all geared up for sample processing

**Fig 3:**
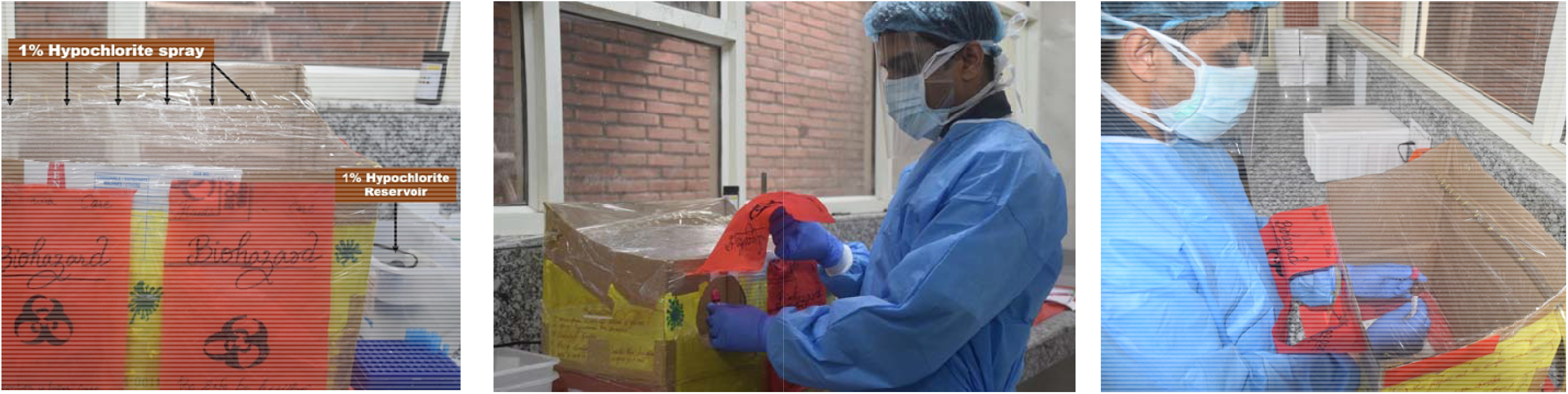
Decapper with sanitization. Demonstration of the steps

## References

1. Sim MR. the COVID-19 pandemic: major risks to healthcare and other workers on the front line. Occupational and Environmental Medicine 2020; 77:2020–282.

2. Bowdle A, Munoz-Price LS. Preventing infection of patients and healthcare workers should be the new normal in the era of novel coronavirus epidemics. Anesthesiology 2020; 132(6):1292–1295.

3. Wang W, Xu Y, Gao R, et al. Detection of SARS-CoV-2 in Different Types of Clinical Specimens. JAMA. 2020; 323(18):1843–1844

4. https://www.cidrap.umn.edu/news-perspective/2020/03/study-covid-19-may-spread-several-different-ways

5. Boone SA, Gerba CP. Significance of Fomites in the Spread of Respiratory and Enteric Viral Disease. Applied and Environmental Microbiology 2007, 73(6):1687–1696.

6. Galbadage T, Peterson BM, Gunasekera RS. Does COVID-19 Spread through Droplets Alone? Front. Public Health 2020; 8:163.

7. Rutter DA, Evans CGT. Aerosol hazards from some clinical laboratory apparatus. B MJ1972;1:594–597

8. https://www.who.int/emergencies/diseases/novel-coronavirus-2019/technical-guidance/laboratory-guidance

9. Lippi G, Adeli K, Ferrari M, Horvath AR, Koch D, Sethi S, Wang CB. Biosafety measures for preventing infection from COVID-19 in clinical laboratories: IFCC Taskforce Recommendations. CCLM 2020. https://doi.org/10.1515/cclm-2020-0633

10. Kampf G, Todt D, Pfaender S, Steinmann E. Persistence of coronaviruses on inanimate surfaces and their inactivation with biocidal agents. J Hosp Infect 2020; 104:2020–51.

11. World Health Organization. (2020). Rational use of personal protective equipment for coronavirus disease (COVID-19) : interim guidance, 27 February 2020. World Health Organization. https://apps.who.int/iris/handle/10665/331215. License: CC BY-NC-SA 3.0 IGO

12. European Centre for Disease Prevention and Control. Infection prevention and control for COVID-19 in healthcare settings – March 2020. ECDC: Stockholm; 2020. https://www.ecdc.europa.eu/sites/default/files/documents/COVID-19-infection-prevention-and-control-healthcare-settings-march-2020.pdf.

13. Centers for Disease Control and Prevention. Interim Laboratory Biosafety Guidelines for Handling and Processing Specimens Associated with Coronavirus Disease 2019 (COVID-19). https://apps.who.int/iris/bitstream/handle/10665/331138/WHQ-WPE-GIH-2020.1-eng.pdf.

14. Guidance and Standard Operating Procedure COVID-19 Virus Testing in NHS Laboratories. https://www.rcpath.org/uploads/assets/90111431-8aca-4614-b06633d07e2a3dd9/Guidance-and-SOP-COVID-19-Testing-NHS-Laboratories.pdf.

15. American Biological Safety Association International (ABSA). Considerations for Handling Potential SARS-CoV-2 Samples. https://absa.org/wp-content/uploads/2020/03/ABSA2020_Covid-19-dr3.pdf.

16. First Affiliated Hospital, Zhejiang University School of Medicine (FAHZU). Handbook of COVID-19 Prevention and Treatment. https://esge.org/documents/Handbook_of_COVID19_Prevention_and_Treatment.pdf.

17. Lippi, G., Adeli, K., Ferrari, M., Horvath, A. R., Koch, D., Sethi, S., & Wang, C. (2020). Biosafety measures for preventing infection from COVID-19 in clinical laboratories: IFCC Taskforce Recommendations, Clinical Chemistry and Laboratory Medicine (CCLM), 58(7), 1053–1062.

18. http://www.maggenome.com/shop/rna-extraction-saliva-cellline-kit/

19. Lu R, Zhao X, Niu P, Yang B, Wu H, et al. Genomic characterisation and epidemiology of 2019 novel coronavirus: implications for virus origins and receptor binding. Lancet. 2020 Jan 30. pii: S0140-6736(20)30251-8. doi:10.1016/S0140-6736(20)30251-8.

20. Chan JF, Yuan S, Kok KH, et al. A familial cluster of pneumonia associated with the 2019 novel coronavirus indicating person-to-person transmission: a study of a family cluster. Lancet. 2020; 395(10223):514–523.

21. Corman VM, Landt O, Kaiser M, Molenkamp R, Meijer A, Chu DK, et al. Detection of 2019 novel coronavirus (2019-nCoV) by real-time RT-PCR. Euro Surveill. 2020 Jan; 25(3):2000045.

22. Yam WC, Chan KH, Poon LL, Guan Y, Yuen KY, Seto WH, Peiris JSM. Evaluation of reverse transcription-PCR assays for rapid diagnosis of severe acute respiratory syndrome associated with a novel coronavirus. J Clin Microbiol. 2003; 41(10):4521–4524.

